# Efficacy of N-163 beta-glucan in beneficially improving biomarkers of relevance to muscle function in patients with muscular dystrophies in a pilot clinical study

**DOI:** 10.1101/2023.07.20.23292982

**Authors:** Kadalraja Raghavan, Thanasekar Sivakumar, Koji Ichiyama, Naoki Yamamoto, Mangaleswaran Balamurugan, Vidyasagar Devaprasad Dedeepiya, Rajappa Senthilkumar, Senthilkumar Preethy, Samuel JK Abraham

## Abstract

**Background:** Muscular dystrophies other than Duchenne muscular dystrophy (DMD) are genetic diseases characterized by increasing muscle weakness, loss of ambulation, and ultimately cardiac and respiratory failure. There are currently no effective therapeutics available. Having demonstrated the efficacy of a N-163 strain of Aureobasidium Pullulans (Neu-REFIX) produced B-1, 3-1,6-Glucan in pre-clinical and clinical studies of Duchenne muscular dystrophy (DMD) earlier, we assessed the effectiveness of this novel Beta glucan in the other muscular dystrophies in the present study.

**Methods:** In this 60-day study, six patients with muscular dystrophies other than DMD consumed one 8g gel of Neu-REFIX beta-glucan along with their usual standard of care treatment regimen, and their biomarkers of relevance to muscle function such as serum calcium (SC), creatinine phosphokinase (CPK), and alkaline phosphatase (ALP) levels along with functional improvement criteria, viz., Medical research council (MRC) scale and North Star Ambulatory assessment (NSAA) were assessed at baseline and following the intervention.

**Results:** After the intervention, the SC levels significantly decreased from a mean baseline value of 9.28 mg/dL to 8.31 mg/dL (p-value = 0.02). With a p-value of 0.29, the mean CPK value dropped from 2192.33 IU/L to 1567.5 IU/L. Following the intervention, the ALP levels dropped from 200.33 to 75.5 U/L (p-value =0.15). MRC scale improved in three out of six patients. NSAA remained stable. There were no adverse effects.

**Conclusion:** This study has proven the safety of Neu REFIX beta-glucan food supplement and its efficacy in improving both plasma biomarkers and functional parameters of muscle in a short duration of 2 months. Further validation by evaluation of muscle function for a longer duration is recommended to confirm the efficacy of Neu-REFIX food supplement as a potential adjuvant DMT in muscular dystrophies.

## Introduction

Muscular dystrophies (MDs) are a group of about 30 distinct hereditary disorders with specific anomalies such as variation in muscle fibre size, muscle fibre necrosis, scar tissue formation, and inflammation. These MDs include dystrophinopathies such as Duchenne muscular dystrophy (DMD), Emery-Dreifuss muscular dystrophies, congenital muscular dystrophies, limb-girdle muscular dystrophies (LGMD), and fascioscapulohumeral muscular dystrophy. The heterogeneity of these different disorders is delineated by a combination of clinical, genetic, molecular, and pathological aspects [1]. Among MDs other than DMD, Limb-girdle muscular dystrophy (LGMD) is common wherein LGMD1 refers to autosomal dominant conditions and LGMD2 to recessive disorders. The sarcoglycan complex plays a key role in the aetiology of LGMD with four types of autosomal recessive LGMD caused by mutations in the sarcoglycan gene. LGMD affects the skeletal muscles, is genetically inherited and causes progressive, primarily proximal muscular weakening due to muscle fibre loss. To be defined as LGMD, the disease has to be present in at least two unrelated families, the affected individuals have to be able to walk independently, the serum creatine kinase (CK) level has to be elevated and the muscle imaging has to show degenerative changes, the muscle histology has to show dystrophic changes, and the disease should eventually progress to end-stage pathology for the most affected muscles [2]. Individual genetic mutations that primarily cause protein shortage or misfolding give rise to the LGMD subtypes. Even though it is unclear how these proteins affect mitochondrial function, it is obvious that they play a part in energy production, maintaining Ca2+ homeostasis, or activating the apoptotic pathway [3]. It is generally accepted that the prevalence of LGMD across all subtypes ranges from 0.8 to 6 per 100,000, For LGMD, both generic and disease-specific therapies are being developed. There are now atleast 25 different therapeutic approaches in various phases of development across all stages of research and commercialization. These treatments include anti-myostatin treatments, modulation of the immune system using steroids, Coenzyme Q10 and lisinopril, gene therapies such as Gamma-sarcoglycan gene-containing recombinant AAV1 vector-based therapy, SRP-9004: Sarcoglycan gene stimulator and small molecule therapies [4] but all have associated side effects and cannot be applied to all patients due to the heterogenous presentation of the disease.

We have previously reported the beneficial effects of a 1-3,1-6 beta-glucan from the N-163 strain of the black yeast Aureobasidium pullulans (Neu REFIX) in decreasing inflammatory biomarkers such as interleukin (IL)-6, tumour growth factor (TGF)-, and IL-13 and increased dystrophin levels as well as improved muscle strength in patients with DMD in a clinical study conducted in 28 patients for a 45-day period [5] and then the muscle improvement in a study of 6-months duration [6]. The gut microbiome was also found to be beneficially reconstituted in the same study [7]. We also performed a pre-clinical study of this N-163 beta-glucan in a mdx mouse model[8], which showed a significant decrease in inflammation score and fibrosis as well as levels of plasma alanine transaminase, aspartate aminotransferase, lactate dehydrogenase, and haptoglobin; increased anti-inflammatory TGF-levels; and balanced regulation of the amount of centrally nucleated fibres indicative of muscle regeneration [8] apart from increase in CD44 and MYH3 expression indicative of muscle regeneration followed by maturation [9]. In the present study we have evaluated the effects of this Neu REFIX B-glucan in patients with MDs other than DMD.

## Materials and methods

This trial was an investigator-initiated, single-centre, open-label, prospective, linear, single-arm clinical study of patients with MDs other than DMD. This study was conducted over a period of 60 days.

The intervention was consumption of one sachet of N-163 beta-glucan (Commercial name: Neu-REFIX) once daily along with their standard of care treatment regimen.

### Inclusion criteria

1. Subjects diagnosed with LGMD and muscular dystrophies other than DMD
2. Male and Female subjects of age in between 3 years to 70 years
3. Subjects and legally authorized representative for vulnerable subjects must be willing to participate in the study and provide a written informed consent
4. Subject willing to and able to comply with the protocol

### Exclusion criteria

1. Patients with a previous (within the past 1 month) or concomitant participation in any other therapeutic trial
2. Subjects and legally authorized representative for vulnerable subjects who has not given informed consent for this study
3. Subjects with history of multiple sclerosis
4. Subjects with history of any muscular atrophy
5. Pregnant and lactating females

### Investigations

Data on sex, date of birth, age, habits, medical history, medications, treatments, allergies (to foods and drugs), regular use of specific foods for health purposes, functional foods, health foods, consumption of foods high in beta-glucan and foods containing beta-glucan, and consumption of immunity-boosting foods were collected as part of a background survey. Height, weight, body mass index, and temperature readings were recorded as well as a medical history. The following measurements and analysis were performed at baseline and at the end of the study (2 months).

1. Levels of IL-6 and Myoglobin urea
2. Levels of Creatinine Kinase, Titin, TNF-Alpha, Haptoglobin, Liver function test, Complete blood count, Cystatin C, ESR
3. The Medical Research Council (MRC) Scale was used to measure muscle strength.
4. North Star Ambulatory Assessment (NSAA)

The participants were contacted every week for drug compliance and the recording of adverse effects, if any.

### Statistical analysis

Microsoft Office Excel statistics package and the software Origin 2021b were used for statistical analysis. Statistical significance was set at p<0.05.

## Results

Six patients, five male and one female were included in the study. The mean ± standard deviation of age for the total study population was 28 ± 16.11 years (range, 8–51 years). Table 1 shows the genetic diagnosis of the subjects.

**Table 1.**

| Subject | Gender | Age (Range)<br>years | Genetic diagnosis |
| --- | --- | --- | --- |
| I | Female | 11-15 | LGMD Type 2E |
| II | Male | 21-25 | Miyoshi MD-1, LGMD Type 2B |
| III | Male | 51-55 | LGMD- Type 1 |
| IV | Male | 16-20 | LGMD- Type 1 |
| V | Male | 41-45 | Nonaka myopathy |
| VI | Male | 6-10 | Clinical diagnosis of MD; No<br>genetic diagnosis |

MD: Muscular Dystrophy; LGMD: Limb Girdle Muscular Dystrophy

No adverse events were observed. No clinically significant changes from baseline data were observed on physical examination or in vital signs.

The serum calcium levels significantly decreased from a mean baseline value of 9.28 ± 0.59 mg/dL to 8.31 0.89 mg/dL post-intervention (p-value = 0.02). The mean CPK value decreased from 2192.33 IU/L to 1567.5 IU/L (p-value =0.29). The ALP levels decreased from 200.33 to 75.5 U/L (p-value =0.15) post-intervention. The TNF-Alpha levels increased from a baseline of 17.95 ± 2.45 to 31.08 ± 25.81 pg/mL (p-value =0.12). There was not any significant changes in the other biochemical parameters measured. The MRC scale improved from 117.6 ± 22.59 to 120 ± 24.51 in three patients while the score decreased from 148.66 ± 2.88 to 142.33 ± 0.57 in the remaining three patients. NSAA remained stable before and at the end of the study.

## Discussion

The goal of treatment approaches to muscular dystrophies (MD) including LGMD has been to focus on improving the quality of life of patients while maintaining mobility and functional independence for as long as possible apart from effectively managing the comorbidities. The complexity of the disease makes the challenges enormous [10]. Any uniformity in strategy is impossible due to high variation in presentation of the disease. The relative scarcity of targeted therapeutics is explained by heterogenous patterns of inheritance, clinical epidemiology, gene abnormalities, protein expression, and pathophysiology. It appears extremely improbable that any one therapy strategy will be effective. Corticosteroids can alter the natural course of DMD by prolonging ambulation by two to three years and supporting the maintenance of pulmonary function but in the case of other MDs such as LGMD, majority of the treatment is supportive and interdisciplinary. Current objectives typically focus on maintaining function rather than improving it [10].

Growth hormone, myoblast transfer, and myostatin inhibition have all failed to ameliorate MDs so far; available and reliable information on pharmacologic methods such as corticosteroids, myostatin inhibition, and myostatin inhibition is sparse. Gene therapies are still under development and have not reached the clinic [10]. So, until a definitive approach is identified, modulating the inflammation and immune response by a safe approach that can be administered to all the patients in an easy manner is needed.

The biological response modifier glucan (BRMG) approach using the Neu REFIX Beta-Glucan has been by far safe and successful in ameliorating the inflammation, fibrosis in not only DMD [5–9] but in multiple sclerosis (MS) [11], non-alcoholic steatohepatitis (NASH) [12] and COVID-19 [13]. In DMD apart from beneficial outcome in biochemical parameters and muscle function in studies of 45 days and 6 months duration, there has been increase in levels of dystrophin which is the major protein that is affected by the gene mutation in DMD and this outcome is usually expected only in gene or exon skipping therapies but not in disease modifying treatments (DMTs) which makes the Neu REFIX Beta-glucan, an effective DMT for muscular dystrophies.

Injury to the plasma membrane (PM) results in an abnormal calcium influx. Through the ER-resident calcium pumps and the calcium-activated ion channels, ER aids in the removal of this increase in cytoplasmic calcium [14]. In order to facilitate calcium import into the ER, ion channel maintains the electroneutrality of the ER lumen by anion import. When one or both of these transporter activities are inhibited, cytosolic calcium excess and PM repair (PMR) are both affected. Patients with MDs other than LGMD, due to disruption of the calcium pumps and ion-channel mechanisms have compromised cytosolic and ER calcium homeostasis. By Addressing calcium overload in these myofibers enables them to repair. In the present study there has been significant decrease in serum calcium levels. Elevated levels of enzymes such as CPK and ALP in MD patients is attributed to muscle damage and leakage [15]. There has been significant decrease in CPK and ALT levels in the present report.

Though TNF-alpha is largely produced by macrophages as an inflammatory response mediator and high circulating TNF-is thought to be a pathogenic component, it is now known that TNF-Alpha is constitutively expressed by myoblasts and that its activity is momentarily elevated in developing myoblasts, therefore highlighting their physiological role in myogenesis and muscle regeneration [16]. In the current study, the increase in TNF-Alpha apart from improvement in MRC grading in 50% of the patients in a short duration of 60 days is worth being considered as a significant clinical improvement.

The smaller sample size, heterogeneity of the subjects, treatment regimens used, and a short duration are the study’s limitations. However, the safety and effectiveness of this N-163-produced beta-glucan dietary supplement in improving muscle function without causing side effects can be viewed as an essential adjunct to disease-modifying therapies for MDs.

## Conclusion

The N-163 Beta-Glucan (Neu-REFIX) is a promising disease-modifying pharmacological adjuvant in MDs other than DMD as it has improved the key biochemical parameters of relevance viz., serum calcium, CPK, ALT and TNF-Alpha apart from improving MRC in 50% of the patients in this short duration study of 60 days. Longer duration multi-centric studies are recommended to validate the potential of this safe, allergen-free, orally consumable food supplement as a DMT adjuvant in slowing the progression of MDs.

## Declarations

### Ethics approval and consent to participate

The study was registered in Clinical trials registry of India, CTRI/2022/05/042917 and approved by the ethics committee of Saravana Multispeciality Hospital-Institutional Ethics Committee, India

### Consent for publication

Not applicable

## Availability of data and material

All data generated or analysed during this study are included in the article itself.

## Authors’ contributions

KR, NI and SA. contributed to conception and design of the study. KR, TS and RS helped in data collection and analysis. SA and SP drafted the manuscript. NY, KI, VD and MB performed critical revision of the manuscript. All authors read and approved the final manuscript.

## Data Availability

All data produced in the present work are contained in the manuscript

## Acknowledgements

The authors thank

1. The Government of Japan and the Prefectural Government of Yamanashi for a special loan and M/s Yamanashi Chuo Bank for processing the transactions.

2. Ms. Sunitha, Mr. Vincent and the staff of JAICARE and Sarvee Integra, Dr. Ragaroobine, Mr. Rajmohan from Nichi-In Centre for Regenerative Medicine (NCRM) for their assistance during the clinical study and data collection of the manuscript.

3. Fr. Francis Xavier, Rector, Loyola College, Chennai, Fr. Vargheesh Antony and Fr. Marianathan of JAICARE for their support during the clinical study.

4. Ms. Misa Takamoto, Mr. Masato Onaka, Mr. Yasushi Onaka of Sophy Inc, Kochi, Japan for necessary technical support.

5. Ms. Yoshiko Amikura and staff of GN Corporation, Japan for their liaison assistance with the conduct of the study.

6. Loyola-ICAM College of Engineering and Technology (LICET) for their support to our research work.

---

**Figure 1:**
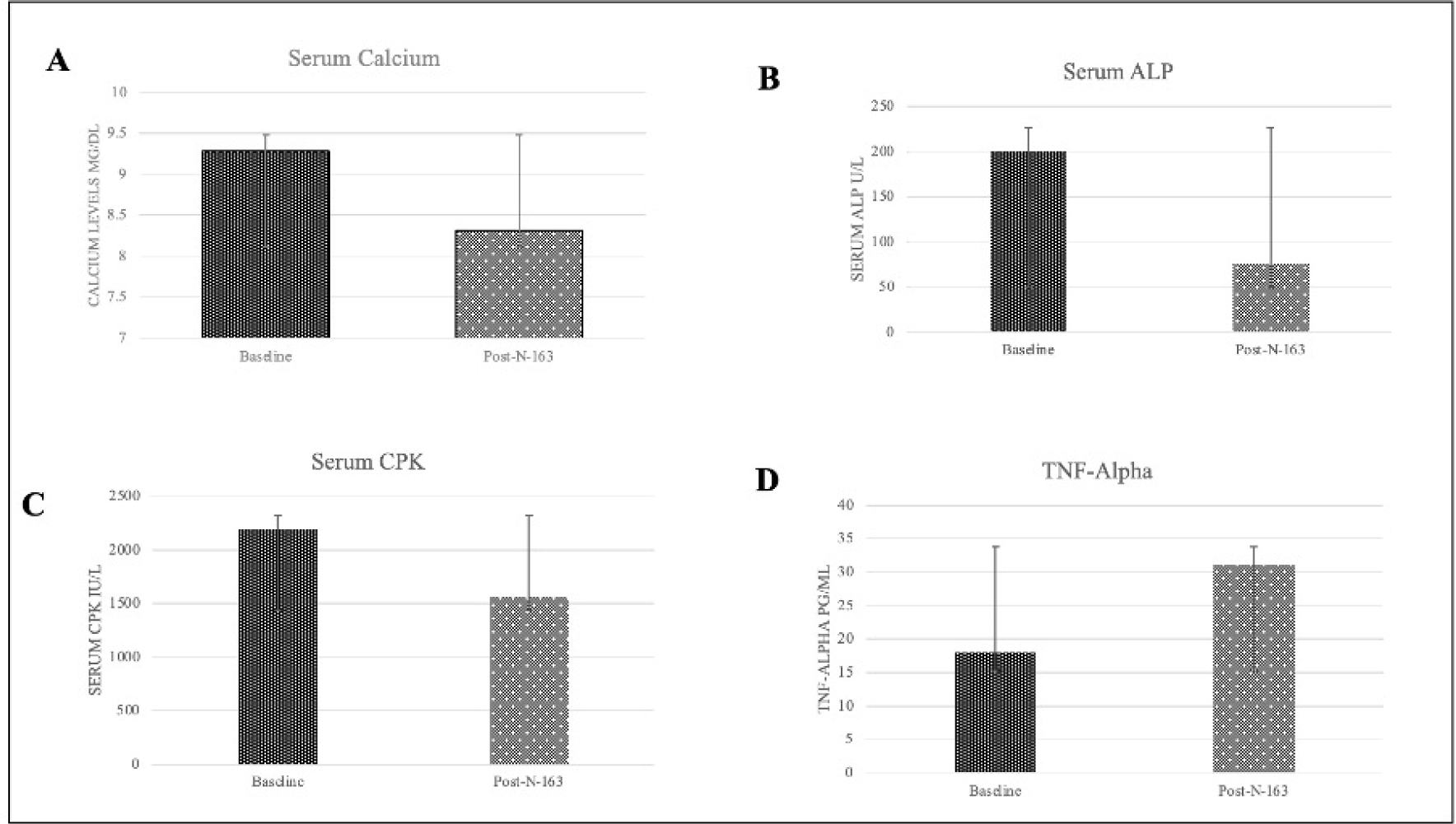
Serum Calcium, Alkaline Phosphatase (ALP), Creatinine phosphokinase (CPK) and Tumour necrosis factor-Alpha in LGMD patients at baseline and 60 days post-intervention with N-163 B-Glucan (Neu REFIX)

**Figure 2:**
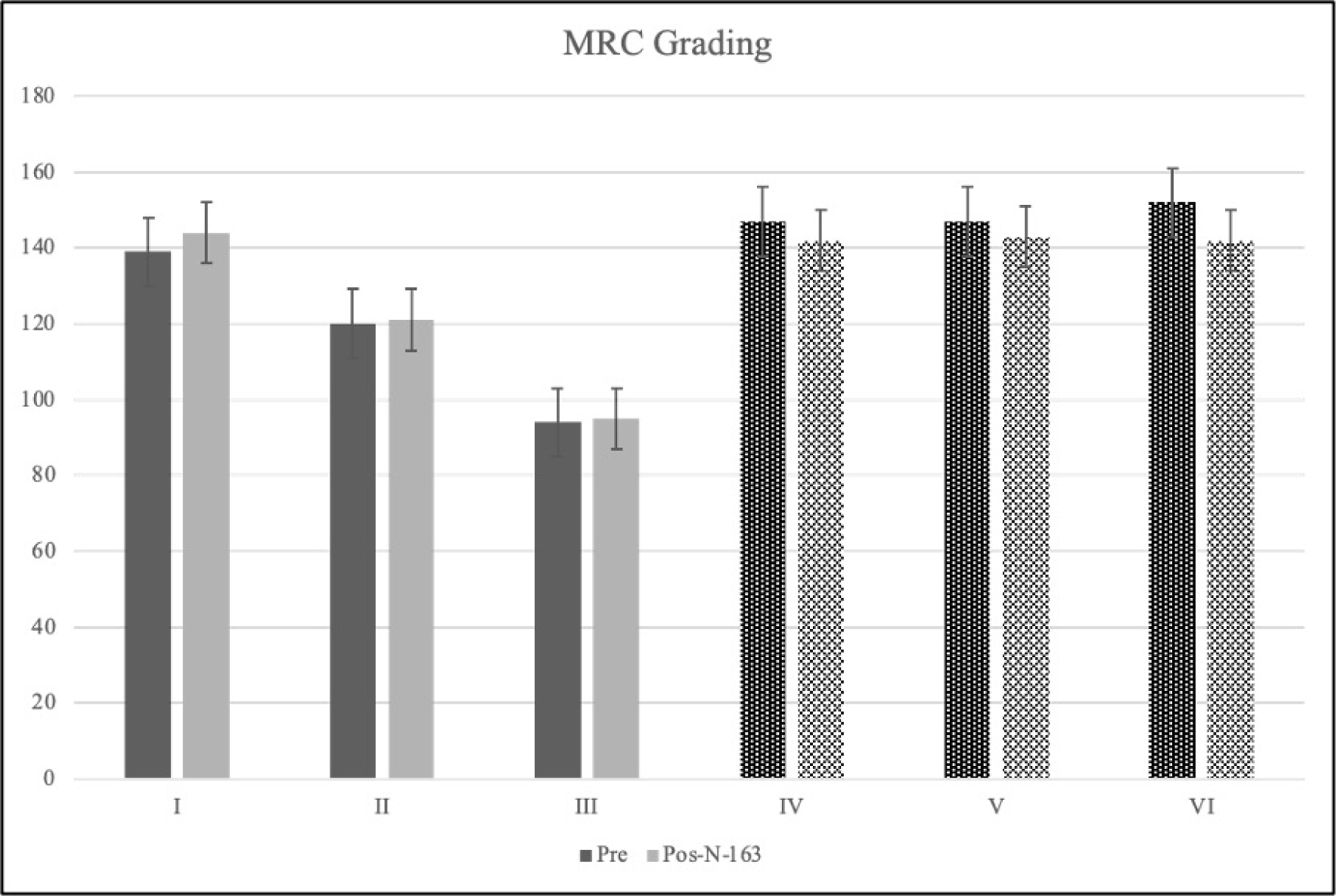
Medical Research Council grading pre- and post-intervention of N-163 beta-glucan in patients with LGMD showing increase in values (improvement) in I-III patients and decrease in IV-VI patients.

